# Detecting Preclinical Alzheimer’s Disease Risk in Cognitively Normal Adults Using Speech Acoustics: Validation with Plasma p-Tau217 and APOE-ε4 Status

**DOI:** 10.64898/2026.01.15.26344226

**Authors:** Mehrdad Dadgostar, Lindsay C. Hanford, Maryam Tavakoli, Steven E. Arnold, David H. Salat, Tatiana Sitnikova, Pia Kivisakk Webb, Jordan R. Green, Hengru Liu, Brian D. Richburg, Mariam Tkeshelashvili, Marziye Eshghi

## Abstract

**INTRODUCTION:** We tested whether spontaneous speech acoustics provide a scalable digital marker of biologically defined Alzheimer’s disease (AD) risk.

**METHODS:** Forty-nine cognitively unimpaired older adults were stratified within APOE genotype into Low-, Moderate-, and High-Risk groups based on log₁₀-transformed plasma p-tau217. Acoustic features were extracted from spontaneous speech and entered into multiclass SVM classifiers with leave-one-out cross-validation, with and without genetic-algorithm feature selection and age. Parallel models using neuropsychological measures were evaluated for comparison. Feature contributions were interpreted using SHAP.

**RESULTS:** Speech-based models substantially outperformed cognition-only models and exceeded chance performance for three-group classification (33.3%), achieving up to 77% accuracy compared with 47% for neuropsychological models. SHAP analyses identified a compact, stage-dependent acoustic signature dominated by voice-quality, spectral-envelope, and formant-bandwidth features, with age contributing secondary effects.

**DISCUSSION:** Spontaneous speech acoustics capture p-tau217/APOE-defined AD risk despite preserved cognition, supporting speech as a scalable, biologically grounded biomarker for preclinical AD risk stratification.

## 1. Background

Despite projections that Alzheimer’s disease (AD) will affect more than 150 million people worldwide by 2050 [1,2], there is still no scalable, clinically implemented strategy to identify individuals at highest risk during the preclinical stage [3]. This stage can extend for more than a decade [4], yet the absence of a validated clinical framework for risk stratification delays opportunities for timely intervention and personalized treatment planning, when disease-modifying interventions would be most impactful [5,6]. Although traditional neuropsychological assessments are considered the gold standard for evaluating cognitive function, they lack the sensitivity to detect the subtle molecular, physiological, and neuropathological changes that precede the emergence of overt symptoms [5,7,8]. As a result, early pathological processes often remain undetected until measurable cognitive decline emerges. At the same time, current biomarker-based tools, including cerebrospinal fluid (CSF) assays and amyloid PET imaging, provide biologically specific information but require specialized infrastructure, entail per-scan costs in the several-thousand-dollar range, and carry procedural risks related to lumbar puncture [9–11]. These barriers limit population-level implementation and exacerbate existing health disparities [1,6], leaving most at-risk individuals unidentified. These limitations highlight the urgent need for scalable, non-invasive, and cost-effective approaches capable of detecting preclinical AD with sensitivity to its earliest biological changes.

One promising, underutilized avenue for preclinical AD detection is speech. Quantifiable speech markers reliably distinguish individuals with mild cognitive impairment (MCI) and AD from cognitively unimpaired adults [12–17], positioning speech as an objective complement to traditional, largely subjective cognitive assessments. However, most work grouped under the broad label of “speech” has centered on linguistic features (e.g., lexical diversity, syntactic complexity, semantic coherence, discourse organization, and pause-filling behavior) that primarily index higher-order cognitive and language processes and typically deteriorate only around or after the onset of MCI. In contrast, speech motor control, captured through acoustic properties (e.g., timing, pitch stability, spectral features, and voice quality) and articulatory kinematics (e.g., movement amplitude, speed, and coordination of the lips, tongue, and jaw), has received far less attention despite its potential sensitivity to early neuromotor changes. Speech production relies on the tightly coordinated interaction of distributed cortical and subcortical networks, so even subtle pathological changes within these systems are plausibly reflected in fine-grained alterations of speech timing, stability, and articulatory precision.

Neuroimaging studies demonstrate that regions supporting speech motor control including precentral and supplementary motor areas show tau-related dysfunction and altered activation patterns across the AD continuum, even in the absence of frank motor symptoms [18,19]. Critically, many of these regions lie within, or are strongly connected to, Braak I–IV networks [20–22] where hyperphosphorylated tau first accumulates and seeds broader network degeneration, disturbing synaptic plasticity, axonal transport, and large-scale connectivity before clinical diagnosis. APOE-ε4 further amplifies this vulnerability by potentiating amyloid–tau interactions and accelerating tau accumulation and spread within these networks, including motor- and language-connected regions [23–25]. Against this mechanistic backdrop, emerging work indicates that subtle alterations in speech acoustics and articulatory kinematics can already be detected in cognitively-intact APOE-ε4 carriers [26], supporting the notion that speech motor metrics may index subclinical vulnerability of these tau-sensitive circuits. These findings support the hypothesis that speech motor metrics represent scalable, non-invasive indicators of preclinical AD risk, with particular sensitivity to molecular and genetic insults that precede clinical symptom onset.

This study tests the hypothesis that speech-derived acoustic features can detect early manifestations of AD pathophysiology in cognitively normal adults at high biological risk, operationalized by elevated plasma phosphorylated tau-217 (p-Tau217) and APOE-ε4 carrier status. Plasma p-Tau217 is a highly sensitive and specific blood-based biomarker that captures both amyloid and tau burden and closely tracks PET-derived measures of AD pathology [27–29]. By centering analyses on p-Tau217, this work directly links speech alterations to underlying disease biology. This biologically anchored strategy positions speech analysis as a promising tool for early AD detection and large-scale risk stratification, moving beyond purely phenomenological digital markers toward mechanistically informed biomarkers. By enabling accessible, scalable, and non-invasive screening in the preclinical stage, this approach could reshape clinical practice by opening a critical window for earlier, targeted intervention in at-risk but asymptomatic individuals.

## 2. Methods

### 2.1. Participant population

The study included 49 adult subjects (28 males and 21 females) with a median age of 68 years (mean 68.48, SD: 7.18; range 57 to 84 years). Venous blood samples were collected from all participants following standardized protocols. Genomic DNA was extracted from peripheral blood leukocytes and genotyping for APOE-ε4 alleles was performed using established polymerase chain reaction (PCR) methods. Plasma concentrations of p-tau217 were quantified employing the FujiRebio Lumipulse G p-tau217 immunoassay platform. A total of 30, participants, all homozygous for the ε3 allele (ε3/ε3), were classified as non-carriers (ε4^−^). An additional 19 participants were identified as APOE-ε4 carriers (ε4^+^), possessing either the heterozygous ε3/ε4 or homozygous ε4/ε4 genotypes. All participants were blinded to both their APOE genotype and p-tau217 plasma concentration throughout the study. Table 1 presents demographic and clinical characteristics stratified by APOE ε4 carrier status. APOE ε4 carriers and non-carriers were generally comparable across most demographic, cognitive, and clinical measures. Sex distribution, family history and Educational did not differ significantly between ε4⁺ and ε4⁻ groups (both P>.05). A significant difference was observed for age (P=.009), with ε4 non-carriers being older on average than ε4 carriers. Despite this age difference, global cognitive performance remained comparable between groups. In addition, APOE ε4 carrier status did not significantly affect performance on executive function and memory measures (Table 1). Neuropsychological tests used in this study are described below.

**Table 1.**
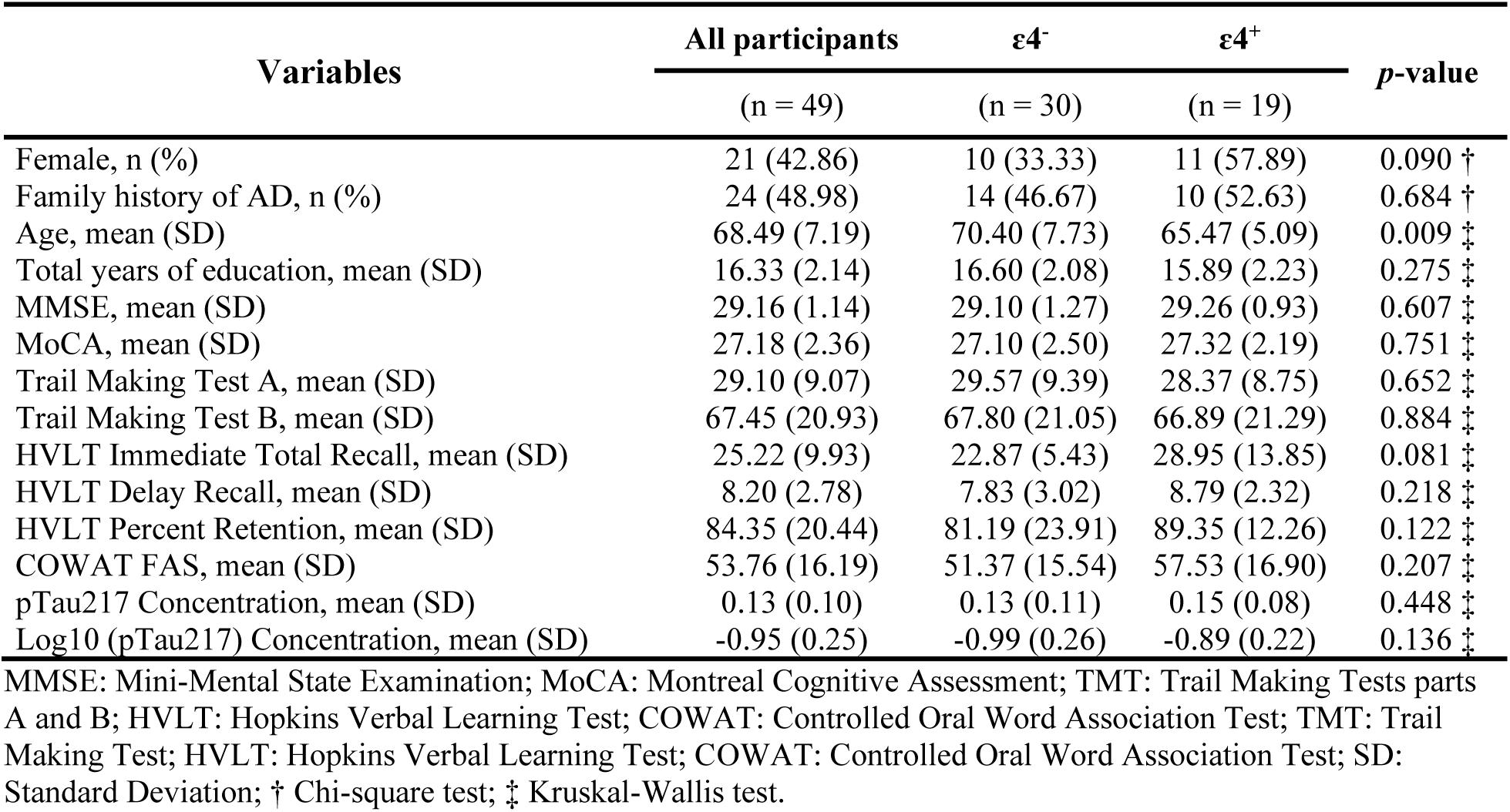
Demographic and clinical characteristics stratified by genotype carrier status.

| Variables | All participants | $\epsilon 4^-$ | $\epsilon 4^+$ | *p*-value |
| --- | --- | --- | --- | --- |
|  | (n = 49) | (n = 30) | (n = 19) |  |
| Female, n (%) | 21 (42.86) | 10 (33.33) | 11 (57.89) | 0.090 † |
| Family history of AD, n (%) | 24 (48.98) | 14 (46.67) | 10 (52.63) | 0.684 † |
| Age, mean (SD) | 68.49 (7.19) | 70.40 (7.73) | 65.47 (5.09) | 0.009 ‡ |
| Total years of education, mean (SD) | 16.33 (2.14) | 16.60 (2.08) | 15.89 (2.23) | 0.275 ‡ |
| MMSE, mean (SD) | 29.16 (1.14) | 29.10 (1.27) | 29.26 (0.93) | 0.607 ‡ |
| MoCA, mean (SD) | 27.18 (2.36) | 27.10 (2.50) | 27.32 (2.19) | 0.751 ‡ |
| Trail Making Test A, mean (SD) | 29.10 (9.07) | 29.57 (9.39) | 28.37 (8.75) | 0.652 ‡ |
| Trail Making Test B, mean (SD) | 67.45 (20.93) | 67.80 (21.05) | 66.89 (21.29) | 0.884 ‡ |
| HVLT Immediate Total Recall, mean (SD) | 25.22 (9.93) | 22.87 (5.43) | 28.95 (13.85) | 0.081 ‡ |
| HVLT Delay Recall, mean (SD) | 8.20 (2.78) | 7.83 (3.02) | 8.79 (2.32) | 0.218 ‡ |
| HVLT Percent Retention, mean (SD) | 84.35 (20.44) | 81.19 (23.91) | 89.35 (12.26) | 0.122 ‡ |
| COWAT FAS, mean (SD) | 53.76 (16.19) | 51.37 (15.54) | 57.53 (16.90) | 0.207 ‡ |
| pTau217 Concentration, mean (SD) | 0.13 (0.10) | 0.13 (0.11) | 0.15 (0.08) | 0.448 ‡ |
| Log10 (pTau217) Concentration, mean (SD) | -0.95 (0.25) | -0.99 (0.26) | -0.89 (0.22) | 0.136 ‡ |

MMSE: Mini-Mental State Examination; MoCA: Montreal Cognitive Assessment; TMT: Trail Making Tests parts A and B; HVLT: Hopkins Verbal Learning Test; COWAT: Controlled Oral Word Association Test; TMT: Trail Making Test; HVLT: Hopkins Verbal Learning Test; COWAT: Controlled Oral Word Association Test; SD: Standard Deviation; † Chi-square test; ‡ Kruskal-Wallis test.

Plasma pTau217 concentrations, both in raw form and following log₁₀ transformation, did not differ significantly between ε4⁺ and ε4⁻ groups (P=.448 and P=.136, respectively). APOE ε4 carrier status alone did not discriminate pTau217 levels in this population, highlighting the need for risk classification based on biomarker thresholds instead of genotype alone.

All participants satisfied the following inclusion criteria: (*i*) capacity to provide informed consent and comply with all study procedures; (*ii*) native fluency in American English; (*iii*) no history of speech, language, hearing, psychiatric, or neurological disorders; (*iv*) absence of a clinical diagnosis of MCI; (*v*) no current use of psychoactive medications that could impact study outcomes; (*vi*) normal age-adjusted hearing acuity in at least one ear, as verified by pure-tone screening; and (*vii*) dental and occlusal status within functional limits, confirmed via oral examination conducted by ME. Additionally, all participants demonstrated normal functional speech, as evaluated by the Sentence Intelligibility Test (SIT) [30], a standardized assessment of speech intelligibility and speaking rate. Each participant read aloud 11 SIT software–generated sentences, ranging in length from 5 to 15 words. Speech samples were digitally recorded and orthographically transcribed by two native American English-speaking raters, who demonstrated high interrater reliability (ICC = 0.99, p < 0.001). SIT-derived metrics included speech intelligibility (percent of accurately transcribed words) and speaking rate (words per minute; WPM). Moreover, participants showed no signs of sound distortion or abnormalities in voice, resonance, prosody, or speech rate, as independently confirmed by two expert raters (ME, JG).

The study was approved by the Mass General Brigham (MGB) Institutional Review Board (IRB Protocol #2021P001460). All procedures were conducted in accordance with MGB IRB guidelines and regulations. Written informed consent was obtained from all participants prior to their participation.

### 2.2. Neuropsychological assessments

Participants completed a comprehensive battery of standardized neuropsychological assessments designed to characterize cognitive profiles associated with AD–related disorders. The battery evaluated multiple domains, including attention, processing speed, learning and memory, language, and executive functioning. The cognitive tests included the Montreal Cognitive Assessment (MoCA) [31–33], Mini-Mental State Examination (MMSE) [34,35], Trail Making Tests (TMT) parts A and B [36,37], Hopkins Verbal Learning Test (HVLT) [38,39] assessing immediate recall, delayed recall, as well as retention efficiency, and the Controlled Oral Word Association Test (COWAT). Test scores were normalized according to established age-and education-adjusted norms to facilitate cross-participant comparisons. A trained neuropsychologists administered and scored all cognitive assessments to ensure standardization and reliability. Test scores were normalized using established age- and education-adjusted norms, and standardized scores were derived for use in subsequent data analyses and classification models.

### 2.3. Experimental tasks

Spontaneous speech samples were collected from all participants. Participants were asked to speak for one minute on each of the following prompts: (1) describe what a typical weekend looks like for you, (2) describe your hobbies and activities, and (3) describe a place or country you have visited and particularly enjoyed. Spontaneous speech tasks are designed to engage natural, unconstrained speech production, capturing a continuum of cognitive–linguistic and speech motor processes, including articulatory coordination, respiratory–phonatory control, and prosodic modulation. These tasks provide ecologically valid insight into how the speech motor system operates in everyday communication and may reveal subtle impairments in the timing, precision, and stability of speech movements that precede overt cognitive or language deficits.

### 2.4. Audio recording

All speech recordings were collected in a sound-attenuated, acoustically treated booth in the Speech Physiology and Neurobiology of Aging and Dementia (SPaN-AD) Laboratory to reduce background noise and reverberation. Participants were seated upright in a comfortable chair, with body position and head orientation standardized relative to the microphone across sessions. Speech was captured using a high-quality, head-mounted unidirectional condenser microphone (Williams AV MIC 094), placed approximately 10–15 cm from the mouth and routed through a Focusrite Scarlett 2i2 interface with gain levels set and checked at the beginning of each visit. Signals were digitized at 22.5 kHz with 32-bit resolution and saved as uncompressed mono WAV files to preserve acoustic detail.

Prior to recording, participants were instructed to speak in their typical voice, at a comfortable loudness and pace, and to minimize non-speech noise (e.g., coughing, chair movement). Each session began with a short calibration utterance to support subsequent level normalization. Recording quality was monitored continuously, and the same seating, microphone position, and hardware settings were maintained for all participants. After each session, recordings were visually and auditorily inspected (waveforms and spectrograms) for artifacts or unexpected noise, and any segments with substantial contamination were re-recorded when feasible.

### 2.5. Data processing and analysis

Digitized audio files were then post-processed in Audacity (Version 3.7.1; Audacity Team) to eliminate extraneous pauses and non-speech segments. Each sample was manually edited to retain only the speech segments corresponding to the target tasks for subsequent analysis.

### 2.6. Statistical analysis

All statistical analyses were conducted using MATLAB (Version 24.2, R2024b; The MathWorks Inc., Natick, MA, USA). APOE-ε4 genotype and plasma p-tau217 measurements were available for all participants. To reduce right-skewness and enhance interpretability of model estimates, plasma p-tau217 values were log₁₀-transformed prior to analysis.

Linear regression analyses were initially performed to characterize the joint effects of age and APOE ε4 genotype on log₁₀-transformed p-tau217 concentrations and to justify the subsequent genotype-specific risk stratification. In these models, age, APOE ε4 status (carrier vs non-carrier), and their interaction were entered as predictors; model assumptions (linearity, homoscedasticity, normality of residuals, and multicollinearity) were evaluated using standard diagnostic procedures, ensuring that the observed age × genotype pattern in p-tau217 was robust and suitable for defining high-, moderate-, and low-risk groups based on within-genotype biomarker distributions.

Subsequently, log₁₀-transformed plasma p-tau217 levels were compared between APOE ε4 carriers (ε4⁺) and non-carriers (ε4⁻) to characterize genotype-related differences in the biomarker distribution. Based on the log₁₀-transformed p-tau217 values within each APOE group, participants were then stratified into three risk categories defined within genotype: High-Risk, comprising individuals in the top quartile (≥75th percentile) of p-tau217 in each APOE group; Moderate-Risk, comprising APOE ε4 carriers below the 75th percentile; and Low-Risk, comprising APOE ε4 non-carriers below the 75th percentile (Figure 1).

**Figure 1.**
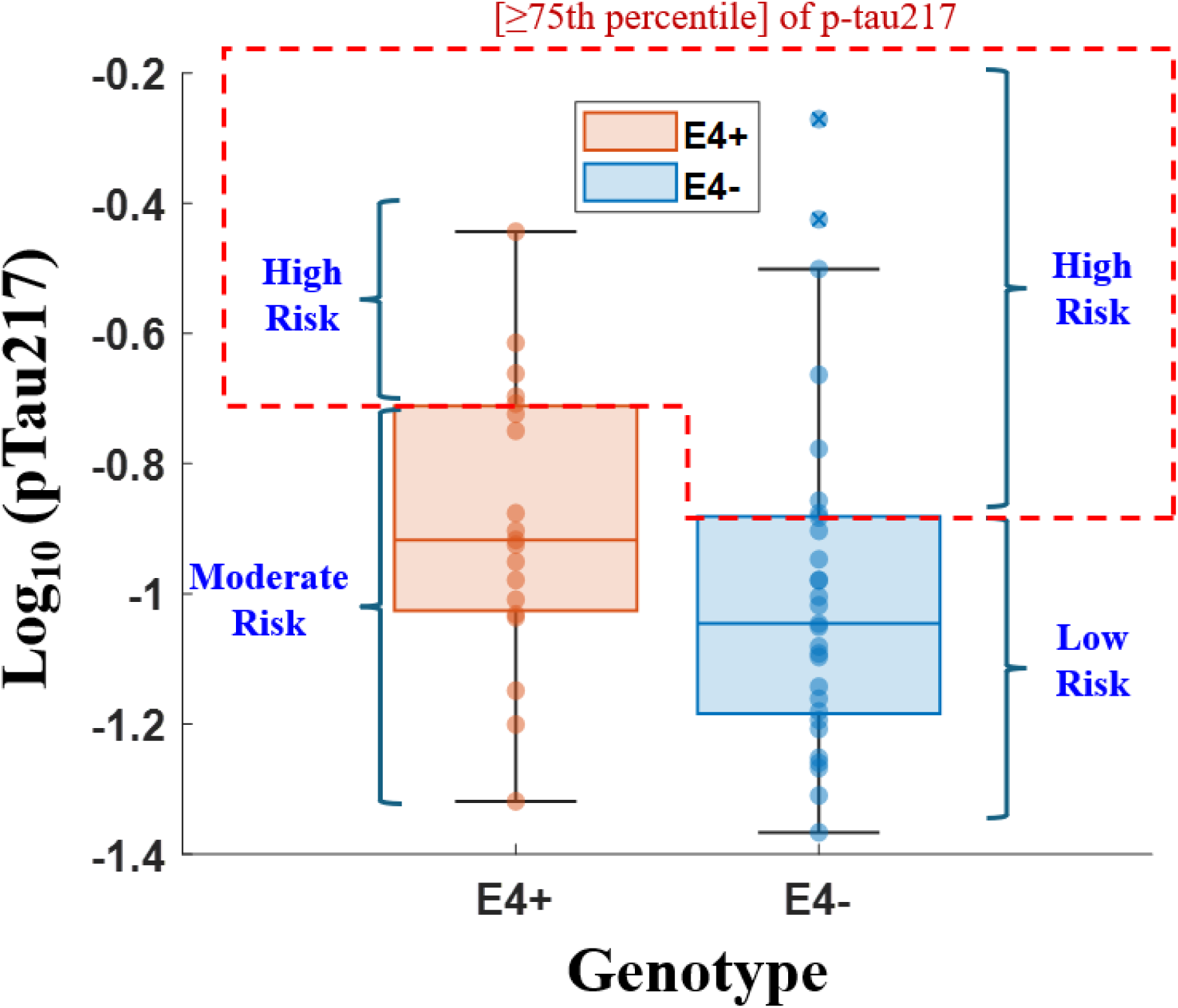
Boxplot of log₁₀-transformed pTau217 levels across APOEε4 genotype groups. There are three risk groups based on APOE genotype: High-Risk (top quartile [≥75th percentile] of p-tau217 in both APOEε4 genotype groups); Moderate-Risk (APOE ε4 carriers below the 75th percentile); and Low-Risk (APOE ε4 non-carriers below the 75th percentile).

Table 2 summarizes the demographic and clinical characteristics of the three risk groups. The groups were generally well balanced with respect to sex distribution and family history of disease, with no significant differences observed (all P>.05). Educational attainment and global cognitive performance, indexed by MMSE and MoCA, were also comparable across Low-, Moderate-, and High-Risk groups, indicating no marked baseline differences in general cognition or educational background. In contrast, age differed significantly across groups (P=.034), with individuals in the High-Risk group being older on average than those in the Moderate- and Low-Risk groups, consistent with an age-related increase in risk classification. Performance on executive function and memory measures, including Trail Making Tests A and B, HVLT immediate and delayed recall, HVLT percent retention, and COWAT FAS, did not significantly differ between groups (all P>.05), indicating preserved and broadly similar neuropsychological profiles at this stage. As expected by design, plasma p-tau217 concentrations showed robust group differences: both raw and log₁₀-transformed p-tau217 values differed significantly across risk groups (both P< .0001), with a graded increase from Low- to Moderate- to High-Risk. This stepwise elevation supports the validity of the stratification scheme and highlights plasma p-tau217 as a sensitive biomarker for distinguishing AD risk levels in the absence of overt cognitive impairment.

**Table 2.** Demographic and clinical characteristics stratified by risk groups.

| Variables | All participants | Low Risk | Moderate Risk | High Risk | *p*-value |
| --- | --- | --- | --- | --- | --- |
|  | (n = 49) | (n = 22) | (n = 14) | (n = 13) |  |
| Female, *n* (%) | 17 (38.64) | 7 (35.00) | 7 (53.85) | 3 (27.27) | 0.371 † |
| Family history of AD, *n* (%) | 22 (50.00) | 9 (45.00) | 8 (61.54) | 5 (45.45) | 0.611 † |
| Age, mean (SD) | 68.34 (7.41) | 69.35 (8.57) | 64.15 (4.36) | 71.45 (6.17) | 0.034 ‡ |
| Total years of education, mean (SD) | 16.25 (2.15) | 16.45 (1.90) | 15.77 (2.17) | 16.45 (2.62) | 0.664 ‡ |
| MMSE, mean (SD) | 29.09 (1.18) | 29.30 (1.26) | 29.08 (1.04) | 28.73 (1.19) | 0.187 ‡ |
| MoCA, mean (SD) | 27.07 (2.35) | 27.35 (1.90) | 26.85 (2.38) | 26.82 (3.12) | 0.871 ‡ |
| Trail Making Test A, mean (SD) | 28.64 (9.12) | 29.05 (10.16) | 27.38 (9.19) | 29.36 (7.58) | 0.720 ‡ |
| Trail Making Test B, mean (SD) | 67.57 (20.25) | 64.55 (20.77) | 66.23 (17.84) | 74.64 (22.03) | 0.604 ‡ |
| HVLT Immediate Total Recall, mean (SD) | 24.91 (10.31) | 22.70 (5.46) | 28.46 (13.55) | 24.73 (12.40) | 0.308 ‡ |
| HVLT Delay Recall, mean (SD) | 7.98 (2.75) | 7.55 (3.25) | 8.85 (2.23) | 7.73 (2.24) | 0.449 ‡ |
| HVLT Percent Retention, mean (SD) | 83.26 (21.07) | 78.10 (27.78) | 90.68 (11.73) | 83.88 (12.37) | 0.308 ‡ |
| COWAT FAS, mean (SD) | 53.41 (16.07) | 52.10 (16.83) | 59.77 (17.66) | 48.27 (10.60) | 0.197 ‡ |
| pTau217 Concentration, mean (SD) | 0.14 (0.10) | 0.08 (0.02) | 0.11 (0.04) | 0.26 (0.12) | < .0001 ‡ |
| Log10 (pTau217) Concentration, mean (SD) | -0.94 (0.25) | -1.10 (0.13) | -0.98 (0.17) | -0.62 (0.19) | < .0001 ‡ |

MMSE: Mini-Mental State Examination; MoCA: Montreal Cognitive Assessment; TMT: Trail Making Tests parts A and B; HVLT: Hopkins Verbal Learning Test; COWAT: Controlled Oral Word Association Test; TMT: Trail Making Test; HVLT: Hopkins Verbal Learning Test; COWAT: Controlled Oral Word Association Test; SD: Standard Deviation; † Chi-square test; ‡ Kruskal-Wallis test.

Group-level differences in age, plasma pTau217 concentration, and log₁₀-transformed pTau217 values were examined using one-way analyses of variance (ANOVA) with risk group (low, moderate, high) as the independent factor. For each outcome variable, we first evaluated overall group effects. When the overall ANOVA was significant or indicated group variability, post-hoc pairwise comparisons were conducted using Tukey’s Honestly Significant Difference (HSD) tests to estimate mean differences between risk groups. Plasma pTau217 values were log₁₀-transformed prior to analysis to reduce right-skewness and better satisfy assumptions of normality and homoscedasticity. Model-derived group means and corresponding standard errors were extracted from the fitted linear models, and notched boxplots were generated to visualize the distribution and median differences across risk groups. All statistical tests were two-sided with an α-level of 0.05.

### 2.7. Feature extraction

Acoustic features were extracted in accordance with the extended Geneva Minimalistic Acoustic Parameter Set (eGeMAPS), a standardized protocol for the systematic characterization of paralinguistic voice parameters [40]. Feature extraction was performed using the openSMILE toolkit (Version 2.5.1), ensuring methodological consistency and comparability across samples. The eGeMAPS feature set encompasses a broad range of acoustic parameters including frequency, energy, spectral, and temporal measures and has been widely adopted in clinical and affective computing research to identify subtle alterations in speech production associated with neurodegenerative and psychiatric conditions.

The 88 eGeMAPS features are constructed from a core set of 18 low-level descriptors (LLDs), including frequency-related measures pitch, jitter and formants, energy/amplitude related parameters shimmer, loudness, harmonics-to-noise ratio (HNR), and spectral descriptors MFCCs, spectral flux, alpha ratio and Hammarberg Index. These LLDs are processed using functionals such as the arithmetic mean and coefficient of variation, resulting in 36 initial parameters. Additional functionals (e.g., percentiles, mean and standard deviation of the slopes) applied to pitch and loudness yield 20 further features. The extended set includes seven additional LLDs, primarily cepstral and dynamic features, and their corresponding functionals, along with aggregated statistics for voiced and unvoiced regions. This results in a total of 88 features per utterance. The compact yet informative nature of eGeMAPS makes it particularly suitable for machine learning pipelines targeting clinical speech-based biomarkers.

### 2.8. Classification

Classification of participants into Low-, Moderate-, and High-Risk groups was performed using a Support Vector Machine (SVM) classifier with a linear kernel implemented within a multiclass error-correcting output codes (ECOC) framework, using a one-vs-one coding design. Linear SVMs (optimized with the SMO algorithm) served as base learners, with the soft-margin penalty parameter (BoxConstraint) set to C=1; final class labels were obtained via loss-weighted decoding of the binary classifier outputs. Because SVM performance is sensitive to feature scaling, all input features were standardized prior to model training. Linear SVMs offer strong generalization in high-dimensional feature spaces by maximizing the margin between classes, and the one-vs-one ECOC scheme further improves robustness by decomposing the multiclass problem into balanced pairwise classifiers. Classification performance was quantified using accuracy, precision, sensitivity, specificity, F1-score, and balanced accuracy, all evaluated with leave-one-out cross-validation.

### 2.9. Feature selection using a Genetic Algorithm (GA)

A genetic algorithm (GA) was then used for feature selection to optimize classification performance while limiting overfitting. In this framework, each candidate solution was encoded as a binary chromosome, where each gene indicated whether a given feature was included (1) or excluded (0) from the subset. The GA evolved a population of candidate feature sets via selection, crossover, and mutation to maximize classification performance, using a population size of 2000 and a maximum of 100 generations to balance exploration of the binary feature space with convergence stability. Initial chromosomes were generated using a uniform binary creation function; scattered crossover was applied to recombine parental solutions across randomly chosen loci, and uniform mutation introduced random bit flips to escape local optima. Parent selection was implemented using tournament selection to maintain selection pressure while preserving high-performing individuals across generations. Four acoustic feature configurations were evaluated: (*i*) the full eGeMAPS set (88 features), (*ii*) eGeMAPS plus age (89 features), (*iii*) GA-selected subsets from eGeMAPS alone, and (*iv*) GA-selected subsets from the combined eGeMAPS + *age* feature pool.

In parallel, eight cognitive measures were used as input features for classification: MoCA, MMSE, TMT Parts A and B, HVLT Immediate Total Recall, HVLT Delayed Recall, HVLT Percent Retention, and the COWAT-FAS. Model performance was assessed under two conditions: (*i*) using the full set of cognitive features and (*ii*) applying GA-based feature selection to derive a reduced cognitive feature subset. Classification in all feature configurations was performed with the SVM framework described above.

### 2.10. Shapley Additive Explanations (SHAP)

SHAP (Shapley Additive Explanations) analysis was used to interpret feature contributions in the final SVM models trained on GA-optimized feature sets for each risk-specific classifier and feature configuration (speech-only and speech+*age*). SHAP values were computed with an interventional linear explanation model, providing local attributions for every observation by quantifying how each feature shifted the prediction relative to the model’s baseline output.

In the SHAP summary plots, features were ranked by mean absolute SHAP value to identify features with the greatest overall impact; the sign of each SHAP value indicated whether a feature moved the prediction toward (positive) or away from (negative) a given risk class, and the point color encoded the underlying feature magnitude (cooler to warmer colors for lower to higher values), such that high feature values on the positive SHAP axis reflected risk-enhancing effects, whereas high values on the negative axis reflected lowering or protective effects.

## 3. Results

### 3.1. Age and APOE ε4 Effects on Plasma p-tau217

Figure 2 shows the relationship between age and log₁₀-transformed plasma p-tau217 levels stratified by APOE genotype. In APOE ε4 carriers, p-tau217 shows a clear positive association with age, as reflected by the positive regression slope and its 95% confidence interval, indicating progressively higher p-tau217 concentrations across the age span. In contrast, APOE ε4 non-carriers exhibit a comparatively flat trajectory, with p-tau217 levels remaining relatively stable across ages.

**Figure 2.**
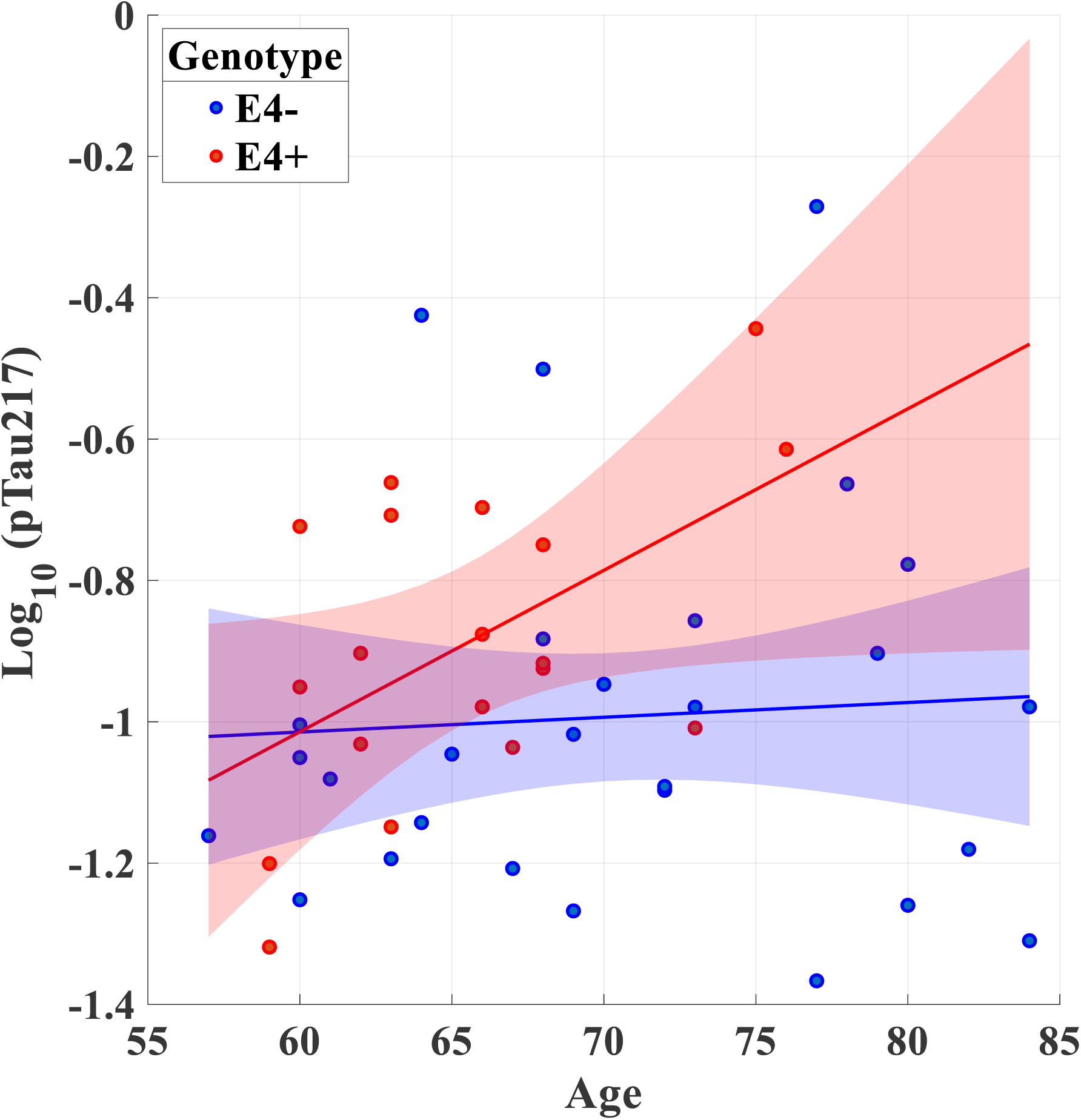
Linear Regression of log₁₀-Transformed plasma pTau217 levels on age, with corresponding 95% confidence intervals for APOE ε4⁺ and APOE ε4⁻ participants.

Linear regression analyses confirmed a significant positive association between age and log₁₀-transformed plasma p-tau217 in APOE ε4 carriers (β = 0.023, p = 0.019, R² = 0.282), consistent with greater tau accumulation with advancing age in this group (Figure 2). No significant association was observed in non-carriers (β = 0.002, p = 0.749, R² = 0.003), indicating minimal age-related change in p-tau217 among ε4⁻ individuals. In the full regression model, age emerged as the only significant feature of p-tau217 levels (p = 0.047), whereas the main effect of genotype (p = 0.146) and the age × genotype interaction (p = 0.106) did not reach statistical significance, although the interaction showed a marginal trend. Together, these findings suggest that age is a primary driver of plasma p-tau217 elevations, with APOE ε4 status potentially shaping the slope of this relationship.

Significant group differences were observed for age and biomarker measures (Table 3). The overall ANOVA for age showed a modest but significant effect of Risk Group (F(2,46) = 3.63, p = 0.034, η² = 0.136), indicating a small-to-medium magnitude of group influence. Although Tukey post-hoc comparisons did not meet the conventional significance threshold, the Moderate-Risk group consistently trended younger than both the Low- and High-Risk groups, a pattern mirrored in model-based mean estimates, with the Moderate-Risk group exhibiting the lowest mean age (Table 3).

**Table 3.** Summary of ANOVA and Post-Hoc Multiple Comparisons Across Risk Groups.

| Variable | Group | Control Group | Risk Group<br>(Mean ± SE) | Lower<br>Limit | Difference | Upper<br>Limit | *p*-value | ANOVA<br>*p*-value |
| --- | --- | --- | --- | --- | --- | --- | --- | --- |
| Age | Low Risk | Moderate Risk | Low: 69.91 ± 1.45 | -0.098 | 5.551 | 11.202 | .055 | .034 |
| | Low Risk | High Risk | Moderate: 64.36 ± 1.82 | -6.411 | -0.629 | 5.152 | .962 | |
| | Moderate Risk | High Risk | High: 70.54 ± 1.89 | -12.547 | -6.181 | 0.184 | .058 | |
| pTau217 | Low Risk | Moderate Risk | Low: 0.079 ± 0.014 | -0.085 | -0.030 | 0.023 | .361 | 1.04$\times 10^{-7}$ |
| | Low Risk | High Risk | Moderate: 0.110 ± 0.017 | -0.224 | -0.169 | -0.113 | 8.28$\times 10^{-9}$ | |
| | Moderate Risk | High Risk | High: 0.248 ± 0.018 | -0.199 | -0.138 | -0.076 | 5.67$\times 10^{-6}$ | |
| Log<sub>10</sub>(pTau217) | Low Risk | Moderate Risk | Low: -1.116 ± 0.033 | -0.261 | -0.133 | -0.004 | .040 | 1.71$\times 10^{-9}$ |
| | Low Risk | High Risk | Moderate: -0.983 ± 0.041 | -0.603 | -0.472 | -0.340 | 8.59$\times 10^{-11}$ | |
| | Moderate Risk | High Risk | High: -0.644 ± 0.043 | -0.483 | -0.339 | -0.194 | 2.65$\times 10^{-6}$ | |

SE: Standard Errors

For plasma p-tau217, the group effect was highly significant (F(2,46) = 28.14, p<1×10⁻⁷, η² = 0.550). Tukey-adjusted pairwise comparisons showed that the High-Risk group had substantially higher p-tau217 concentrations than both Low- and Moderate-Risk groups (all p<.0001), whereas Low-versus Moderate-Risk did not differ significantly. Model-based means demonstrated a clear monotonic increase in p-tau217 across risk groups from Low to High. Analysis of log₁₀-transformed p-tau217 values yielded an even stronger group effect (F(2,46) = 38.15, p<2×10⁻⁹, η² = 0.624). Post-hoc comparisons confirmed significantly elevated transformed p-tau217 levels in the High-Risk group relative to both Low- and Moderate-Risk groups (all p<.0001), and the Low-versus Moderate-Risk comparison reached statistical significance (p=.040). Model-derived means highlighted a pronounced stepwise increase in p-tau217, both raw and log-transformed, with higher risk classification.

### 3.2. Classification Performance using GA

Stratification of participants into risk groups based on APOE genotype and plasma p-tau217 levels successfully delineated high-, moderate-, and low-risk categories for subsequent speech-based classification. In the acoustic models, the baseline SVM using all 88 eGeMAPS features achieved a mean accuracy of 62.69%, exceeding chance performance for a three-class classification. Adding age to the feature set (89 features total) improved mean accuracy to 68.14%. Feature selection via a GA further enhanced performance: GA-optimized eGeMAPS features yielded a mean accuracy of 72.82%, and the combination of GA-selected features with age produced the best results, with a mean accuracy of 76.69% and balanced accuracy above 83% across all risk groups. Across models, precision, sensitivity, specificity, and F1-scores were highest in the age-augmented, GA-optimized configurations, and classification of High-Risk individuals was particularly robust, with sensitivity ≥62.65% and specificity up to 90.47%, indicating strong model capability for identifying those at greatest biological risk (Table 4).

**Table 4.** The results of the classification with different sets. The value in feature set denotes the total number of acoustic features selected.

| Feature set | Group risk | Accuracy (%) | Precision (%) | Sensitivity (%) | Specificity (%) | F1 score (%) | Balanced accuracy (%) |
| --- | --- | --- | --- | --- | --- | --- | --- |
| eGeMAPS (88) | Low risk | 62.69 | 63.84 | 64.96 | 81.40 | 64.40 | 73.18 |
|  | Moderate risk |  | 54.23 | 66.38 | 80.86 | 59.69 | 73.62 |
|  | High risk |  | 62.04 | 62.65 | 81.87 | 62.34 | 72.26 |
|  | Mean |  | **58.14** | **64.51** | **81.36** | **61.02** | **72.94** |
| eGeMAPS + age (89) | Low risk | 68.14 | 64.87 | 69.30 | 81.96 | 67.01 | 75.63 |
|  | Moderate risk |  | 67.25 | 72.60 | 85.91 | 69.82 | 79.26 |
|  | High risk |  | 65.78 | 66.60 | 83.75 | 66.19 | 75.18 |
|  | Mean |  | **66.51** | **69.60** | **84.83** | **67.01** | **75.63** |
| GA-selected eGeMAPS feature set | Low risk | 72.82 | 75.36 | 75.42 | 90.06 | 75.39 | 82.74 |
|  | Moderate risk |  | 57.43 | 81.77 | 83.73 | 67.47 | 82.75 |
|  | High risk |  | 74.32 | 75.48 | 90.47 | 74.89 | 82.97 |
|  | Mean |  | **69.03** | **77.55** | **88.09** | **72.58** | **82.82** |
| GA-selected multimodal (eGeMAPS + age) feature set | Low risk | 76.69 | 76.95 | 81.17 | 90.14 | 77.32 | 85.65 |
|  | Moderate risk |  | 73.01 | 80.19 | 87.81 | 74.23 | 84.00 |
|  | High risk |  | 72.80 | 78.19 | 87.96 | 73.61 | 83.08 |
|  | Mean |  | **72.91** | **79.19** | **87.89** | **73.92** | **83.54** |
| Cognitive features (8) | Low risk | 46.44 | 36.40 | 43.47 | 70.18 | 39.62 | 56.82 |
|  | Moderate risk |  | 27.60 | 32.28 | 72.72 | 29.7 | 52.49 |
|  | High risk |  | 33.20 | 38.85 | 73.24 | 35.80 | 56.04 |
|  | Mean |  | **32.40** | **38.20** | **72.05** | **35.06** | **55.12** |
| GA-selected cognitive feature set | Low risk | 48.44 | 38.00 | 49.71 | 73.12 | 43.07 | 61.41 |
|  | Moderate risk |  | 28.00 | 47.62 | 73.15 | 35.26 | 60.38 |
|  | High risk |  | 34.00 | 47.71 | 76.43 | 39.71 | 62.07 |
|  | Mean |  | **33.33** | **48.35** | **74.23** | **43.07** | **61.41** |

Overall, these findings indicate that speech-derived acoustic features, especially when optimally selected and combined with demographic information such as age, can reliably discriminate among biomarker-defined risk categories and may function as effective, non-invasive markers of preclinical AD risk in research settings.

Figure 3 depicts the distribution of GA-selected features across the full eGeMAPS space and the expanded set including age. Columns represent individual acoustic features grouped by domain (i.e., F0, formants 1–3, spectral measures, MFCCs, loudness, voice quality, and segment duration), while rows correspond to the two GA configurations (eGeMAPS only vs eGeMAPS + *age*). Colored cells indicate features retained by the GA, whereas blank cells denote non-selected features; the gray cell marked with an “X” indicates that age was not part of the candidate feature pool in the eGeMAPS-only GA run.

**Figure 3.**
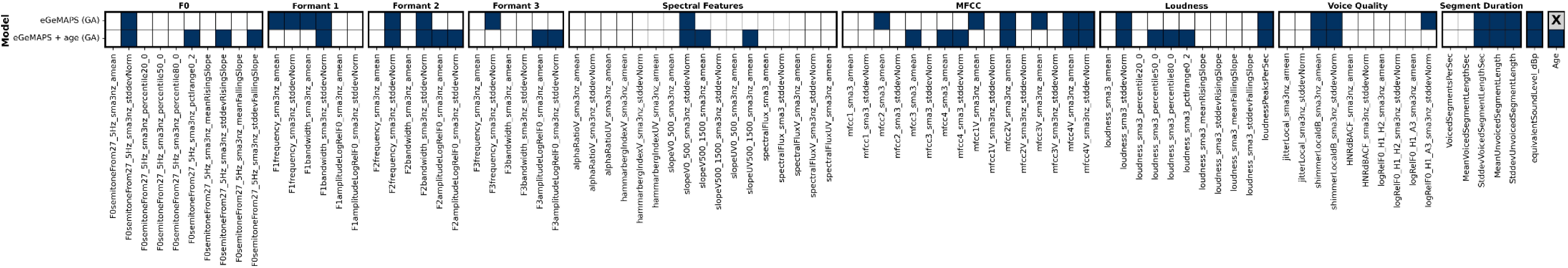
List of features that GA was selected from eGeMAPS (top row) and eGeMAPS+age features (bottom row). The blue-colored cells indicate the features that were selected with GA. Gray cells marked with an ‘X’ denote that the feature Age was not included in the initial set of candidate features.

Genetic algorithm–based feature selection revealed a distributed pattern of acoustic parameters. Overall, the GA selected a sparse and structured subset of features rather than uniformly sampling across acoustic domains, indicating preferential retention of features carrying stable discriminative information across models. Across both GA configurations (i.e., eGeMAPS alone and eGeMAPS combined with *age*) the algorithm consistently retained 16 features (blue cells in Figure 3), reflecting a stable and robust subset of speech markers that recurred despite changes in the candidate feature space. The reproducibility of this overlap underscores the robustness of the selected markers to modifications in feature composition and model specification.

MFCC-based spectral features showed the highest concentration of retained features, highlighting the importance of spectral envelope characteristics and cepstral stability for risk-group discrimination. Segment duration features were also consistently selected, indicating that temporal organization and speech rhythm provide complementary information beyond spectral content alone. In addition, several formant-related variability measures were repeatedly selected, suggesting that instability in vocal-tract resonance patterns contributes meaningfully to model performance. Notably, the overall feature-selection profile was highly similar between two models, indicating that inclusion of age did not substantially alter the core acoustic feature set. Collectively, these findings suggest that accurate risk-group classification arises from the integration of complementary spectral, voice-quality, loudness, and temporal speech attributes rather than reliance on a single dominant acoustic domain.

### 3.3. Cognitive test–based classification performance

Using conventional neuropsychological measures as features yielded modest classification performance across risk groups. Models were trained using MoCA, MMSE, Trail Making Test A and B, HVLT Immediate Total Recall, HVLT Delayed Recall, HVLT Percent Retention, and COWAT (FAS) as features. The initial model achieved a mean accuracy of 46.64%, with precision of 36.40% (Low-Risk), 27.60% (Moderate-Risk), and 33.20% (High-Risk), and sensitivities of 43.46%, 32.27%, and 38.85% for Low-, Moderate-, and High-Risk groups, respectively. Specificity values were 70.17% (Low-Risk), 72.72% (Moderate-Risk), and 73.23% (High-Risk), and F1-scores were 39.62%, 29.70%, and 35.80% for Low-, Moderate-, and High-Risk groups, respectively. Balanced accuracy was 56.82%, 52.49%, and 56.04% for the Low-, Moderate-, and High-Risk groups, indicating only modest discrimination across categories.

Applying a genetic algorithm to select informative cognitive features led to a slight improvement, with mean accuracy increasing to 48.44%. In this optimized model, precision was 38.00% (Low-Risk), 28.00% (Moderate-Risk), and 34.00% (High-Risk), and sensitivity improved to 49.70%, 47.61%, and 47.71% for Low-, Moderate-, and High-Risk groups, respectively. Specificity reached 73.11% (Low-Risk), 73.14% (Moderate-Risk), and 76.43% (High-Risk), with F1-scores of 43.07%, 35.26%, and 39.71% for the three groups. The most discriminative cognitive features identified by the genetic algorithm were MoCA, HVLT Delayed Recall, and COWAT (FAS), suggesting that global cognition, episodic memory consolidation, and phonemic fluency provide the greatest contribution to risk-group classification among the tested measures.

### 3.4. Feature Contributions by SHAP Analysis (Speech-Only Model)

Figure 4 presents SHAP summary plots for the GA-selected eGeMAPS features that explain 95% of the cumulative Shapley importance across the three risk-group classifiers (left: Low-Risk, middle: Moderate-Risk, right: High-Risk). Within each panel, features are ordered by decreasing mean absolute SHAP value, highlighting those with the greatest influence on the classifier’s decision function. Each point corresponds to an individual observation, where the horizontal position (SHAP value) indicates the direction and magnitude of the feature’s contribution relative to the model baseline, and color encodes the underlying feature value (cooler to warmer colors indicating lower to higher values). These summaries illustrate how distinct subsets of acoustic features shape predictions across the three risk-specific models, revealing both shared and group-specific patterns of feature importance and directional effects that support biomarker-defined risk classification.

**Figure 4.**
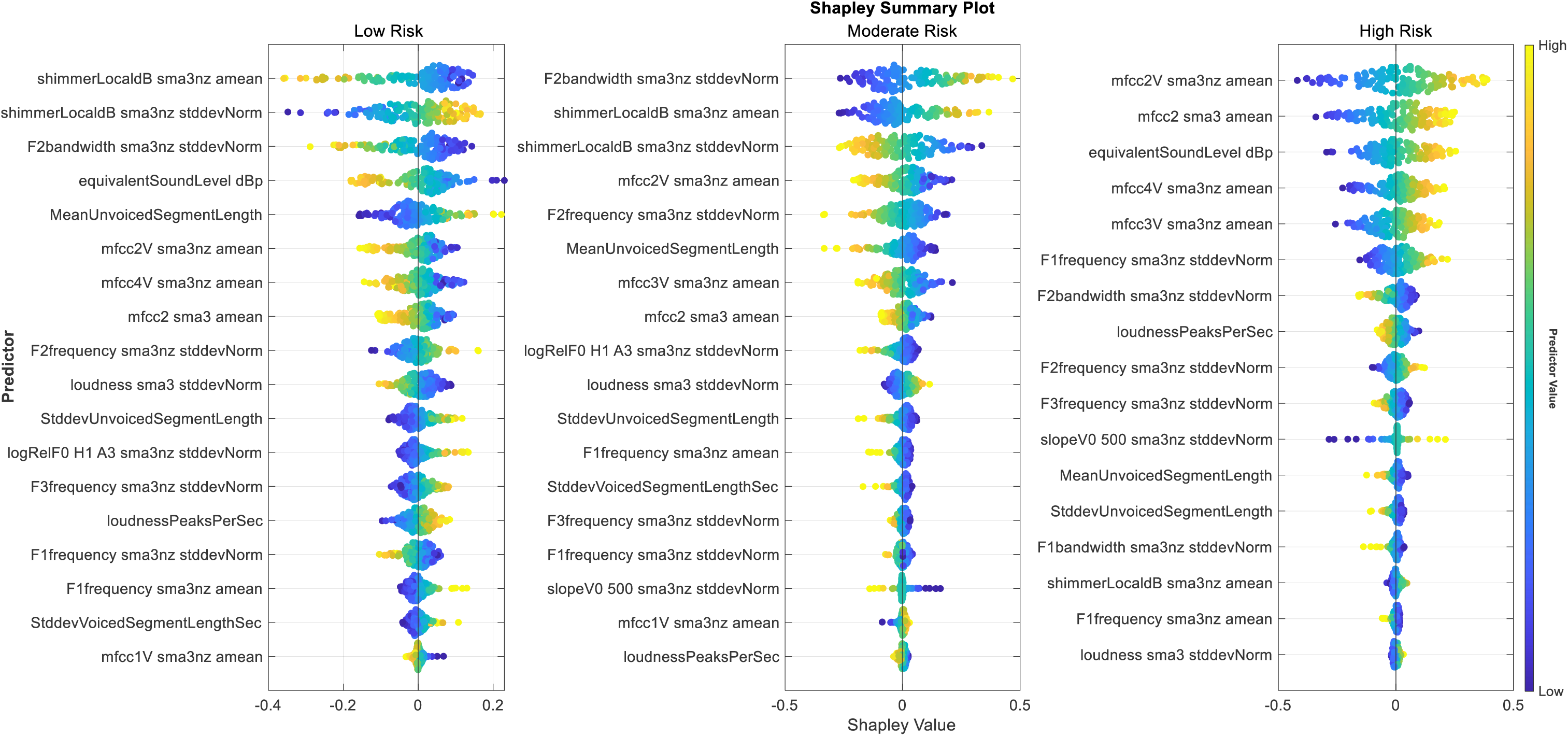
SHAP summary plots of GA-selected eGeMAPS features that explain 95% of the cumulative Shapley importance for risk-group classification, highlighting differential feature importance and directionality across risk groups.

Among voice-quality measures, *shimmerLocaldB_sma3nz*_*amean* and *shimmerLocaldB_sma3nz_stddevNorm* emerged as particularly informative for distinguishing Low-from Moderate-Risk groups. In the Low-Risk classifier, both shimmer features showed strong contributions, with higher shimmer variability (warmer colors) predominantly associated with negative SHAP values, indicating that increased short-term amplitude perturbation pushes predictions away from the Low-Risk class; whereas, lower variability is associated with positive SHAP values, supporting Low-Risk assignment and consistent with preserved vocal stability. In contrast, the Moderate-Risk classifier exhibited a reversal in SHAP directionality for shimmer variability. For *shimmerLocaldB*_sma3nz_*stddevNorm*, higher values tended to cluster on the negative SHAP axis, whereas lower values were more often associated with positive SHAP contributions in the Moderate-Risk classifier. The relative importance of shimmer features was substantially reduced in the High-Risk model, where they ranked below the top 14 features and contributed minimally to classification, and the opposite directional effects observed between the Low- and Moderate-Risk classifiers highlight distinct patterns of amplitude perturbation across earlier stages of risk.

In the High-Risk classifier (right panel of Figure 4), *mfcc2V_sma3nz*_*amean* showed predominantly high-valued observations (yellow or warmer points) with positive SHAP values, indicating that elevated MFCC energy shifted predictions toward the High-Risk class, whereas lower values (blue or cooler points) clustered on the negative SHAP axis and were associated with reduced risk. The wide horizontal spread of SHAP values for this feature underscores its strong contribution to risk-group discrimination, and its directional effect in the High-Risk model is opposite to that observed in the Low- and Moderate-Risk classifiers.

Additionally, *shimmerLocaldB_sma3nz_stddevNorm* in the Moderate-Risk classifier (middle panel) displayed a clear reversal in SHAP directionality as a function of feature magnitude: higher shimmer variability (warmer colors) was predominantly associated with negative SHAP values, indicating that increased short-term amplitude perturbation shifts predictions away from the Moderate-Risk class, whereas lower variability (cooler colors) clusters on the positive SHAP axis, increasing the likelihood of Moderate-Risk classification. This inverse color–direction pattern suggests that Moderate-Risk individuals are characterized by relatively preserved amplitude stability compared with both Low- and High-Risk groups, and that shifts toward higher shimmer variability may instead signal transition toward more advanced impairment; the broad spread of SHAP values further highlights shimmer variability as a key voice-quality marker at this intermediate risk stage.

Across all classifiers, voice quality and spectral stability measures appear to dominate feature importance, with recurrent contributions from shimmer-based descriptors, MFCCs, and formant variability metrics. In the eGeMAPS framework, voice-quality features capture micro-instabilities in phonation such as cycle-to-cycle variations in frequency and amplitude and in the harmonic–noise balance, whereas spectral stability metrics characterize the temporal consistency of the spectral envelope and formant structure, reflecting articulatory precision and motor coordination.

MFCC-based features (*mfcc2V_sma3nz_amean, mfcc3V_sma3nz_amean*, and *mfcc4V_sma3nz_amean*) are described appropriately as having substantial SHAP magnitudes and as reflecting spectral-envelope characteristics related to articulatory and phonatory control. The statement that *mfcc3V_sma3nz_amean* and *mfcc4V_sma3nz_amean* contribute most strongly in the High-Risk model, with weaker influence in the Low- (*mfcc3V_sma3nz_amean*) and Moderate-Risk (*mfcc4V_sma3nz_amean*) classifiers, and that these two features show opposite SHAP directionality across specific pairings of risk groups (i.e., *mfcc4V_sma3nz_amean* between High- and Low-Risk, *mfcc3V_sma3nz_amean* between High- and Moderate-Risk) is conceptually consistent and accurately framed as indicating distinct spectral-envelope alterations that differentially drive classification across risk strata.

Formant-related variability measures, including *F2bandwidth_sma3nz_stddevNorm*, are appropriately described as showing strong but oppositely directed SHAP contributions in the Low-versus Moderate-Risk classifiers, indicating distinct patterns of second-formant bandwidth variability across early risk stages. The interpretation that increased variability in formant bandwidth reflects instability in vocal-tract resonance patterns and suggests altered articulatory control associated with elevated risk status is conceptually sound and consistent with standard acoustic–phonetic principles.

### 3.5. Feature Contributions by SHAP Analysis (Speech + *age* Model)

Figure 5 illustrates SHAP summary plots for the GA-selected multimodal (eGeMAPS + *age*) feature sets that explain 95% of the cumulative Shapley importance across the three risk-group classifiers (Low-, Moderate-, and High-Risk). Across panels, age, voice-quality measures indexing phonatory stability, and *mfcc2V_sma3nz_amean* spectral-stability metrics reflecting articulatory–acoustic consistency dominate feature importance, underscoring their central role in risk-group discrimination.

**Figure 5.**
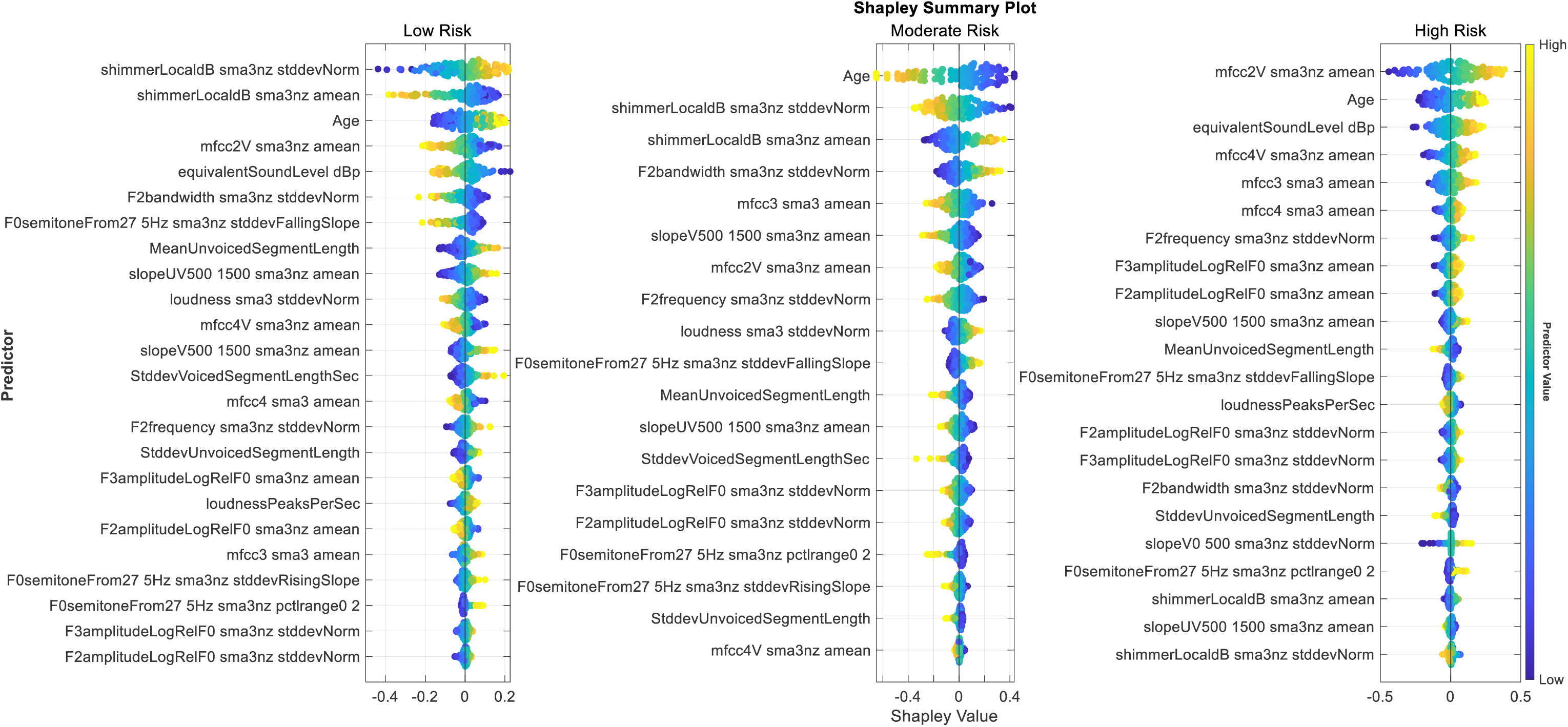
SHAP summary plots of GA-selected multimodal (eGeMAPS + age) features that explain 95% of the cumulative Shapley importance for risk-group classification, highlighting differential feature importance and directionality across risk groups.

In the Low-Risk classifier, the shimmer-related feature (*shimmerLocaldB_sma3nz_stddevNorm*) shows lower feature values (cooler colors) clustering on the negative SHAP axis, indicating that decreased shimmer variability shifts predictions away from the Low-Risk class, whereas higher shimmer variability is associated with positive SHAP values and supports Low-Risk classification consistent with preserved vocal stability. For *shimmerLocaldB_sma3nz_amean*, which ranks second in importance, higher shimmer values are likewise predominantly located on the negative SHAP axis and lower levels on the positive side, reinforcing that amplitude stability reflected by lower mean is characteristic of the Low-Risk group, whereas elevated shimmer tends to push predictions toward higher-risk classes.

In the Moderate-Risk classifier, *shimmerLocaldB_sma3nz_stddevNorm* shows a clear reversal in SHAP directionality relative to feature magnitude: higher shimmer variability (warmer colors) is associated with negative SHAP values, whereas lower variability clusters on the positive SHAP axis. This pattern suggests that Moderate-Risk individuals are characterized by relatively preserved amplitude stability, with increased shimmer variability signaling deviation toward other risk states and contributing less strongly to maintaining Moderate-Risk predictions.

In the High-Risk classifier, *shimmerLocaldB_sma3nz_stddevNorm* contributes modest but consistently across the feature range, with substantial overlap of SHAP values, indicating that shimmer variability alone is not a strong unidirectional discriminator.

In the High-Risk classifier, spectral-envelope features such as *mfcc3V_sma3nz_amean* and *mfcc4V_sma3nz_amean* tend to show larger SHAP magnitudes and more frequent positive SHAP values at higher feature levels, indicating that increased MFCC magnitudes often drive predictions toward the High-Risk class, whereas shimmer-based features have lower rank and smaller SHAP magnitudes, suggesting that spectral alterations play a more prominent role than amplitude perturbation at higher risk levels rather than implying a complete shift away from voice-quality contributions.

In the High-Risk group, spectral envelope features such as *mfcc3V_sma3nz_amean* and *mfcc4V_sma3nz_amean* tend to show larger SHAP magnitudes and more frequent positive SHAP values at higher feature levels, indicating that increased MFCC magnitudes often drive predictions toward the High-Risk class. In contrast, shimmer-based features have lower rank and smaller SHAP magnitudes in this model, suggesting that spectral envelope alterations play a more prominent role than amplitude perturbation in classification at higher risk levels rather than implying a complete shift away from voice-quality contributions.

Age appears as an influential feature across the SHAP summary plots, but its impact is relatively modest and context-dependent compared with the dominant acoustic features. The sign and magnitude of age-related SHAP values indicate how increasing age shifts model predictions toward or away from a given risk group after accounting for all other features: lower ages (cooler colors) tend to be associated with negative SHAP values in the Low- and High-Risk classifiers, while lower ages tend to align with positive SHAP values in the Moderate-Risk classifier, supporting Moderate-Risk assignment.

Across risk groups, age emerged as an influential but clearly secondary predictor relative to dominant acoustic features. In the Low-Risk classifier, age showed modest SHAP magnitudes with mixed directionality, indicating a limited and context-dependent contribution that did not dominate risk assignment. In the Moderate-Risk classifier, age ranked higher than in the Low-Risk model and exhibited a clearer SHAP spread, suggesting increased relevance at this transitional stage. In the High-Risk classifier, age again contributed meaningfully but with narrower SHAP ranges than MFCC-based spectral-envelope features. While the models appropriately capture age-associated risk, classification decisions are driven mainly by speech-derived biomarkers, demonstrating that risk stratification is not dominated by chronological aging but instead reflects pathology-related acoustic changes.

Overall, these findings demonstrate risk-stage–specific patterns of acoustic contribution, with amplitude-stability features most discriminative at early and intermediate stages and spectral-envelope alterations increasingly dominating classification at higher risk. SHAP interpretation of the multiclass ECOC-SVM models therefore provides mechanistic insight into the GA-optimized speech-based classifiers, indicating that voice-quality measures reflecting micro-instabilities in phonation and spectral-stability measures indexing temporal consistency of the spectral envelope jointly shape ECOC decision boundaries and modulate model confidence in multiclass risk discrimination.

## 4. Discussion

The present study demonstrates that speech acoustics offer a powerful, scalable, and non-invasive tool for preclinical AD risk stratification grounded in molecular pathology rather than clinical symptoms. By linking speech motor behavior to p-tau217– and APOE-ε4–defined risk, the work bridges molecular mechanisms of AD and neuromotor control, validating subtle acoustic alterations as early indicators of neurodegenerative vulnerability rather than late manifestations of cognitive decline. Beyond diagnostic applications, these findings support speech as an accessible modality for monitoring biological risk trajectories and therapeutic response in presymptomatic populations, laying the groundwork for precision prevention strategies in AD.

### 4.1. Multidomain Acoustic Signatures

Across GA configurations, feature selection consistently identified a compact but stable acoustic signature dominated by temporal measures, formant- and bandwidth-related parameters, spectral features, and MFCC-derived descriptors. This multidomain feature set captured variability in energy, phonation, articulation, and temporal speech organization, aligning with established neurodegenerative mechanisms involving motor planning, neuromuscular control, and sensorimotor integration, and echoing prior reports that combinations of temporal, spectral, and formant features are sensitive to MCI and AD [41–46]. The consistency of these selections across models indicates that early neuromotor disruption manifests as coordinated changes across multiple acoustic domains rather than isolated abnormalities in a single parameter, providing a physiologically grounded representation of speech production impairments that formed the basis for subsequent classification and interpretability analyses.

### 4.2. Core acoustic feature set across models

Analysis of the GA-selected feature sets showed that risk-group classification relied on a subset of acoustic features spanning voice quality, spectral–formant variability, cepstral dynamics, and temporal speech organization, with several features consistently retained across independent GA runs. A core group of features including *shimmerLocaldB_sma3nz_amean*, *shimmerLocaldB_sma3nz_stddevNorm*, *mfcc2V_sma3nz_amean*, *mfcc2_sma3_amean*, *mfcc4V_sma3nz_amean* and *F2bandwidth_sma3nz_stddevNorm* remained highly ranked and displayed similar SHAP directionality in both speech-only and speech+*age* models, indicating robust contributions to distinguishing Low-, Moderate-, and High-Risk groups that are primarily sensitive to biomarker-defined AD risk rather than chronological aging. This stability suggests that these spectral, formant, and voice-quality markers act as pathology-linked, age-robust digital biomarkers, effectively “de-sensitizing” the classifiers to age while retaining strong sensitivity to p-tau217 and APOE-ε4 status.

### 4.3. Stage-dependent SHAP patterns across risk

The SHAP results reveal that not only the importance but also the direction of key acoustic features shifts systematically from Low- to Moderate- to High-Risk, consistent with stage-dependent reorganization of speech motor control. In the Low-Risk classifier, lower shimmer (mean and variability) and relatively stable formant bandwidths are associated with positive SHAP values and thus favor Low-Risk assignment, indicating that preserved amplitude and resonance stability characterize individuals with minimal biomarker burden. As risk increases to the Moderate group, shimmer variability and F2 bandwidth variability begin to show reversals in SHAP directionality, with certain ranges of reduced variability now supporting Moderate-Risk predictions, suggesting a constrained, “stiffened” speech motor pattern rather than frank degradation.

By the High-Risk stage, the dominant direction of change shifts toward spectral-envelope features: higher MFCC magnitudes (for example, *mfcc2V_sma3nz_amean*, *mfcc2_sma3_amean*, *mfcc3V_sma3nz*_*amean* and *mfcc4V_sma3nz_amean*) carry strongly positive SHAP values and drive predictions toward High Risk, whereas lower MFCC values align with negative SHAP values and reduced risk. In this context, shimmer-based features contribute less and lose their earlier directional asymmetry, implying that once global spectral and articulatory breakdown emerges, localized amplitude perturbations become relatively less informative for discrimination. Across the continuum, these directional flips map a trajectory from “beneficial” stability (low shimmer and modest spectral variability supporting Low Risk), through a transitional regime where reduced variability in some domains marks Moderate Risk, to a high-risk pattern dominated by exaggerated spectral-envelope changes, reflecting a mechanistic gradient from subtle neuromotor dysregulation to overt articulatory–phonatory degradation.

These stage-wise shifts show that speech does not change in a simple linear “more impairment with more risk” fashion but instead follows a patterned reconfiguration of motor control strategies across the risk continuum. At low risk, stability in amplitude and resonance appears protective; at moderate risk, constrained variability and altered bandwidth dynamics emerge as distinct intermediate signatures; and at high risk, global spectral-envelope degradation becomes the dominant driver of classification, suggesting a progression from subtle, potentially compensatory adjustments in phonatory and articulatory control to loss of flexibility in resonance patterns and ultimately overt spectral breakdown.

This staged pattern is consistent with broader neurodegenerative literature showing that early Alzheimer’s and related conditions are characterized by subtle reductions in articulatory agility and precision, which progress toward more global breakdown of speech motor control as disease advances. Studies of connected speech and articulatory kinematics in MCI and dementia report slower, less variable articulation rates, reduced prosodic modulation, and diminished articulatory precision, all interpreted as loss of flexibility in motor planning and execution rather than simple noise [26,47,48]. These observations lend external support to the notion that the SHAP-derived trajectory from stable, flexible control at low risk through constrained, “stiffened” patterns at moderate risk to pronounced spectral and articulatory degradation at high risk captures a mechanistically meaningful gradient of evolving speech-motor impairment across the AD risk continuum.

### 4.4. Stage-specific acoustic profiles and age effects

The SHAP analysis provided an interpretable view of how specific acoustic features and age contributed to multiclass risk-group classification. MFCC-based spectral-envelope measures, voice-quality indices, and formant-variability metrics were consistently ranked among the most influential features across Low-, Moderate-, and High-Risk classifiers, underscoring the central role of fine-grained phonatory and articulatory control in speech-based risk categorization. In the High-Risk group, MFCC features showed the largest SHAP magnitudes, whereas shimmer-based measures were most discriminative at lower and intermediate risk stages and formant-bandwidth variability contributed most strongly at Moderate Risk, reinforcing the notion of stage-specific dominance of different acoustic systems.

Age emerged as an influential but secondary feature, with relatively narrow SHAP ranges compared with dominant acoustic features and context-dependent effects across risk groups. This pattern indicates that the models capture age-related risk while remaining primarily driven by speech-derived biomarkers and that the core acoustic features function as pathology-sensitive, age-robust markers rather than simple reflections of healthy aging.

### 4.5. Interaction of p-tau217, genotype, age, and speech

The study found a significant positive association between age and log₁₀-transformed plasma p-tau217 among APOE-ε4 carriers, but not non-carriers, suggesting that ε4 accelerates age-related tau accumulation. This pattern converges with extensive evidence that APOE-ε4 amplifies age-dependent tau burden and neurodegeneration in preclinical and prodromal AD [25,49,50]. Although the genotype × age interaction did not reach conventional significance in the full model, the observed trend aligns with prior evidence that APOE-ε4 exacerbates tau-mediated neurodegeneration in preclinical and prodromal phases, and the finding that the Moderate-Risk group was youngest despite intermediate p-tau217 levels underscores a dissociation between biological risk and chronological age. Both raw and log-transformed p-tau217 measures showed large effect sizes and a clear monotonic increase from Low- to High-Risk groups, with effect magnitudes exceeding those observed for age, reinforcing p-tau217 as a highly sensitive index of AD pathological burden in this cohort.

This biological stratification provides a plausible mechanistic bridge between molecular pathology and speech. Cortical tau aggregation prominently involves association and premotor cortices that support speech motor planning and higher-order motor control [51–53], suggesting that tau-related disruption of these networks may manifest as the multidomain acoustic changes quantified here. Converging evidence that p-tau217 relates to speech timing and pause measures further supports speech as an integrative biomarker reflecting the combined effects of molecular pathology, genetic susceptibility, and age-related neurobiological changes [54,55]. By anchoring speech-based acoustic measures to p-tau217- and APOE-defined risk groups, the present work strengthens the biological interpretability of digital speech biomarkers and supports their potential for distinguishing normal from pathological aging trajectories.

### 4.6. Cognitive versus speech-based risk classification

The findings indicate that conventional neuropsychological measures provide only modest discriminative power for biomarker-defined AD risk compared with speech-based models. Classification using MoCA, MMSE, Trail Making A/B, HVLT indices, and COWAT (FAS) yielded mean accuracies below 50% and only moderate balanced accuracies, even after GA-based feature selection. In contrast, SVM models trained on GA-selected acoustic features achieved mean accuracies above 70% and balanced accuracies exceeding 83% when age was added, with particularly robust sensitivity and specificity for identifying High-Risk individuals. This divergence suggests that, at preclinical and early-risk stages defined by p-tau217 and APOE-ε4, speech acoustics capture subtle neuromotor changes that are not yet reflected in standard cognitive scores, which often remain near ceiling.

This contrast underscores the clinical value of speech as a complementary, biologically informed digital biomarker that can flag elevated AD risk while cognition is still ostensibly intact. By outperforming cognitive-test–based models in a p-tau217/APOE-defined framework, speech-based classification offers a scalable means to enrich preclinical trial cohorts, prioritize individuals for confirmatory biomarker testing, and monitor subtle disease progression or treatment response long before traditional neuropsychological measures become abnormal.

## 5. Limitations

Several limitations should be considered when interpreting these findings. First, the sample size was modest and drawn from a single cohort of cognitively normal older adults, which may constrain statistical power and limit generalizability despite the use of cross-validation and genetic-algorithm–based feature reduction; replication in larger, independent, and preferably multi-site cohorts is therefore essential. Second, risk groups were defined using plasma p-tau217 thresholds within APOE genotype strata, a biologically motivated but cohort-specific strategy that may introduce threshold effects and should not be equated with definitive clinical disease states. Third, age differed across risk groups and, although explicitly modeled and partially attenuated in SHAP analyses, residual confounding by age or other demographic factors cannot be fully excluded. Future work should therefore prioritize longitudinal, multi-site studies that combine speech recordings with comprehensive biomarker panels including PET and CSF measures, to test whether the identified acoustic signatures predict subsequent cognitive decline, conversion to symptomatic stages, and treatment response. It will also be important to evaluate the robustness of the core feature set across tasks, recording devices, languages, and analytic pipelines, and to integrate speech-derived markers into multimodal risk scores alongside other low-burden digital and clinical measures. Such efforts will clarify the generalizability, causal ordering, and practical deployment of speech-based biomarkers within preclinical AD screening and precision-prevention frameworks.

## Consent Statement

Recruitment and consent procedures adhered to Mass General Brigham Healthcare (MGH) and HIPAA regulations. The study was approved by the MGB Institutional Review Board (IRB Protocol #2021P001460).

Before participating in testing, administrators thoroughly explained the experimental tasks and procedures, addressing any questions participants had. Relevant consent forms were provided, and each form was reviewed point by point with potential participants by a research staff member prior to obtaining their signature. Participants were explicitly informed of their right to withdraw from the study at any time during the experiment without any consequences and all participants provided written informed consent prior to participation.

## Data Availability

The raw speech recordings generated during the current study contain personally identifiable information and cannot be shared publicly for privacy and ethical reasons. A de-identified dataset comprising the extracted acoustic features that support the findings of this study is available from the corresponding author upon reasonable request.

## Author Contribution

M.D. contributed to data analysis, statistical methodology, and manuscript preparation. L.C.H. reviewed the manuscript and provided feedback. M.T. reviewed the manuscript and provided feedback. S.E.A. was involved in participant characterization and reviewed and provided feedback on the manuscript. D.H.S. contributed to participant characterization, manuscript review, and feedback. T.S. reviewed the manuscript and provided feedback. P.K.W. reviewed the manuscript and provided feedback. J.R.G. reviewed the manuscript and provided feedback. H.L. reviewed the manuscript and provided feedback. B.D.R. supported data collection, contributed to IRB submission and approval, reviewed the manuscript, and provided feedback. M.Tk. assisted with post-processing of audio signals for acoustic analysis. M.E. played a leading role in study conceptualization and design, data collection, data analysis, interpretation of findings, and manuscript writing. All authors contributed to the article and approved the submitted version.

